# Trends and Disparities in Respiratory Failure Mortality with Co-Listed Atrial Fibrillation and Flutter Among U.S. Adults Aged ≥65 Years, 1999–2024, with AAMR Forecasts Through 2035: A CDC Wonder Analysis

**DOI:** 10.64898/2026.10.15.26365608

**Authors:** Wareesha Seher, Hafiz Ahmad Masood, Ayesha Jamil, Sakeena Fiza, Abdullah Jehangir, Rabia Hafeez, Muhammad Arbab, Rania Ahmad, Hameed Ullah, Som Prakash Sharma

## Abstract

**Background:** Respiratory failure (RF) and atrial fibrillation/flutter (AF/AFL) often occur together in older adults, but long-term national mortality trends for the two conditions combined have not been described. We examined trends and disparities in deaths with co-listed RF and AF/AFL among U.S. adults aged 65 years and older from 1999 to 2024, and projected rates through 2035.

**Methodology:** We used the CDC WONDER Multiple Cause of Death database to identify deaths with ICD 10 codes J96 (RF) and I48 (AF/AFL) listed together. Age-adjusted mortality rates (AAMRs) per 100,000 were standardized to the 2000 U.S. population and stratified by sex, race/ethnicity, census region, urbanization, state, and place of death. Joinpoint regression was used to estimate annual percent changes (APCs) and average annual percent changes (AAPCs). Forecasts for 2025 to 2035 were generated with an exponential smoothing (ETS) model.

**Results:** A total of 433,598 deaths were recorded. The overall AAMR rose from 12.8 in 1999 to 66.7 in 2024 (AAPC 8.11%; 95% CI, 7.35 to 8.87; P < 0.0001). Three segments were identified: an increase from 1999 to 2018 (APC 7.73%), a sharp rise from 2018 to 2021 (APC 18.10%), and a decline from 2021 to 2024 (APC -5.25%). Males had a higher AAMR and a steeper increase than females (AAPC 8.44% vs 7.81%). Non-Hispanic White adults had the highest mean AAMR, Hispanic adults had the largest relative increase (AAPC 8.72%), and Non-Hispanic Asian/Pacific Islander adults had the smallest (AAPC 6.23%). The Midwest had the fastest regional growth (AAPC 9.35%). From 1999 to 2020, non-metropolitan areas increased faster than metropolitan areas (AAPC 10.09% vs 8.21%). State AAMRs ranged from 24.7 in Ohio to 85.1 in Louisiana. Most deaths occurred in hospital inpatient settings (64.1%), followed by nursing homes (14.1%) and the home (11.9%). The model projects an AAMR of 122.96 per 100,000 by 2035 (95% prediction interval, 85.42 to 160.50).

**Conclusions:** Mortality involving RF with AF/AFL among older U.S. adults increased more than fivefold over 25 years, with a marked surge during the COVID 19 period and only a partial decline afterward. Differences by sex, race/ethnicity, region, and urbanization point to groups that may benefit from earlier detection and coordinated cardiopulmonary care.

## 1. Introduction

Respiratory failure is a major cause of morbidity and mortality, with increasing AAMR from 3.71 in 1999 to 10.50 in 2023 in older adults^(1)^. Atrial Flutter/Fibrillation is also a major cause of death in the USA, causing about 1,307,809 deaths during the 1999 to 2024 period^(2)^. Both conditions are a major burden for healthcare in the US, with increasing prevalence of AF/AFL^(3)^. A study shows that no of deaths from AF/AFL increased from 117,038.00 in 1990 to 315,336.80 in 2019, and the number is expected to be 404,593.40 by 2029^(4)^.

Respiratory failure and AF/AFL have a complex pathophysiological relationship. It has been found that progressive respiratory insufficiency causes conditions like metabolic abnormalities, myocardial stress, and Atrial arrythmias^(5)^. AF/AFL can occur as a complication of respiratory distress, and the patient may require both mechanical ventilation and antiarrhythmic treatment^(6)^. Likewise, AF/AFL have also been proven to be a risk factor for rapid respiratory response leading to pulmonary oedema and eventually respiratory failure^(7)^. It has also been noted that controlling AF in patients improved respiratory distress in clinical settings^(8)^.

Although previous studies have examined mortality trends in AF/AFL and respiratory failure independently, no study has examined how mortality trends have changed in patients with respiratory failure and AF/AFL together using CDC WONDER (Centers for Disease Control and Prevention Wide-Ranging Online Data for Epidemiologic Research) database. Given the bidirectional relationship between pulmonary and cardiovascular system, understanding long-term mortality patterns in patients with coexisting respiratory failure and AF/AFL is necessary for future stratification and to improve healthcare strategies.

Therefore, this study aims to analyze trends in respiratory failure and AF/AFL related mortality among U.S. adults aged 65 years and older from 1999 to 2024, utilizing the CDC WONDER database as the diseases have more prevalence in old adults. Our study aims in highlighting disparities in demographic factors like race, sex, age and urbanization and providing opportunities in early detection, prevention and targeted interventions to optimize clinical outcomes.

## 2. Methodology

### 2.1 Study Setting and Population

This retrospective study utilized the Centers for Disease Control and Prevention Wide-Ranging Online Data for Epidemiologic Research (CDC WONDER) database to retrieve Multiple Cause-of-Death records from 1999 to 2024 for individuals aged ≥65 years^(9,10)^. Cases of respiratory failure (RF) with comorbid atrial fibrillation/flutter (AF/AFL) were identified using International Classification of Diseases, Tenth Revision (ICD-10) codes J96 and I48, respectively^(11)^. Data were extracted from the 1999–2020 and 2018–2024 Multiple Cause of Death datasets.

### 2.2 Data Extraction

Demographic factors extracted from death certificates included sex (male, female), US Census Bureau region (Northeast, Midwest, South, West)^(12)^, and race/ethnicity (Hispanic, Non-Hispanic (NH) White, NH Black, NH Asian/Pacific Islander, NH American Indian/Alaska Native^(10)^. Geographic urbanization was stratified via the National Center for Health Statistics Urban-Rural Classification Scheme into urban (large metropolitan; population >1 million), medium/small metropolitan (50,000– 999,999), and rural (<50,000) categories^(13)^.

### 2.3 Statistical Analysis

Crude and age-adjusted mortality rates (AAMRs) per 100,000 population with 95% confidence intervals (CIs) were calculated annually and stratified by demographic subcategories. AAMRs were standardized to the 2000 US standard population.

Temporal trends were evaluated using the Joinpoint Regression Program (Version 5.0.2, National Cancer Institute) to calculate Annual Percent Changes (APCs) and Average Annual Percentage Changes (AAPCs)^(14)^. The program fitted log-linear models to identify significant inflection points where the trend slope differed significantly from zero. Statistical significance was determined using two-tailed *t*-tests with a threshold of *P* < .05. Time-series forecasting was performed in Microsoft Excel (Microsoft 365) via the automated Forecast Sheet utility, utilizing the Exponential Smoothing (ETS) algorithm. Based on annual AAMR trends from 1999 to 2024, the ETS model projected point estimates across the 2025–2035 forecasting horizon. Upper and lower bounds were calculated using a 95% prediction interval to quantify temporal uncertainty.

### 2.4 Ethical Considerations

Institutional review board approval and informed consent were waived because this study exclusively utilized publicly accessible, fully de-identified aggregate federal data. The study adheres to the Strengthening the Reporting of Observational Studies in Epidemiology (STROBE) guidelines.

## 3. Results

### 3.1 Overall Trends

Between 1999 and 2024, a total of 433,598 deaths were recorded due to RF with comorbid AF/AFL. The overall AAMR increased significantly from 12.8 per 100,000 population in 1999 to 66.7 in 2024, representing an AAPC of 8.11% (95% CI: 7.35– 8.89; *P* < .0001). Joinpoint regression identified two inflection points (2018 and 2021), yielding three distinct temporal segments: a progressive increase during 1999–2018 (APC = 7.73%), a marked acceleration during 2018–2021 (APC = 18.10%), and a subsequent decline during 2021–2024 (APC = −5.25%).

#### Forecasting analysis

Forecasting analysis conducting using ETS model projected a continued increase in the overall age-adjusted mortality rate (AAMR), reaching 122.96 deaths per 100,000 population by 2035 (95% prediction interval: 85.42– 160.50).

**Figure 1.**
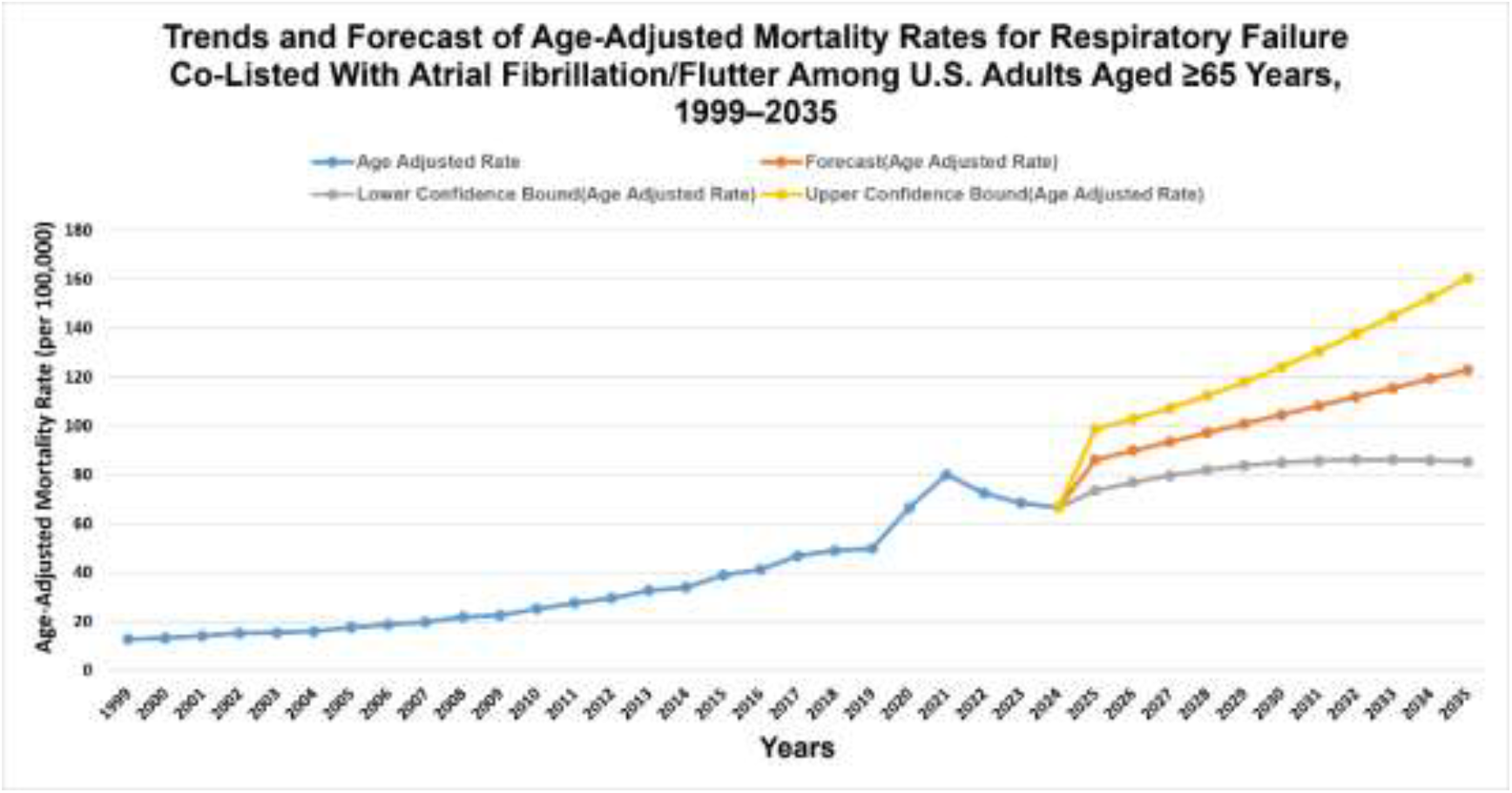
Mortality Trends by Demographic Characteristics (A) Overall mortality trends and Forecast.

### 3.2 Mortality Trends Stratified by Sex

Among males, the AAMR rose significantly from 15.2 in 1999 to 78.7 in 2024. Among females, the AAMR increased from 11.4 in 1999 to 57.8 in 2024. Males demonstrated a higher overall mortality burden and steeper increase than females (overall AAPC: 8.44% vs 7.81%). Among males, the AAMR rise is characterized by a baseline increase during 1999–2018 (APC = 7.56%), a sharp surge during 2018–2021 (APC = 21.16%), and a pronounced post-2021 decline (APC = −7.33%). Females had comparable initial increase in AAMR during 1999–2018 (APC = 7.59%), a less severe pandemic acceleration during 2018–2021 (APC = 15.28%), and a milder subsequent decline during 2021–2024 (APC = −3.48%).

**Figure 1a.**
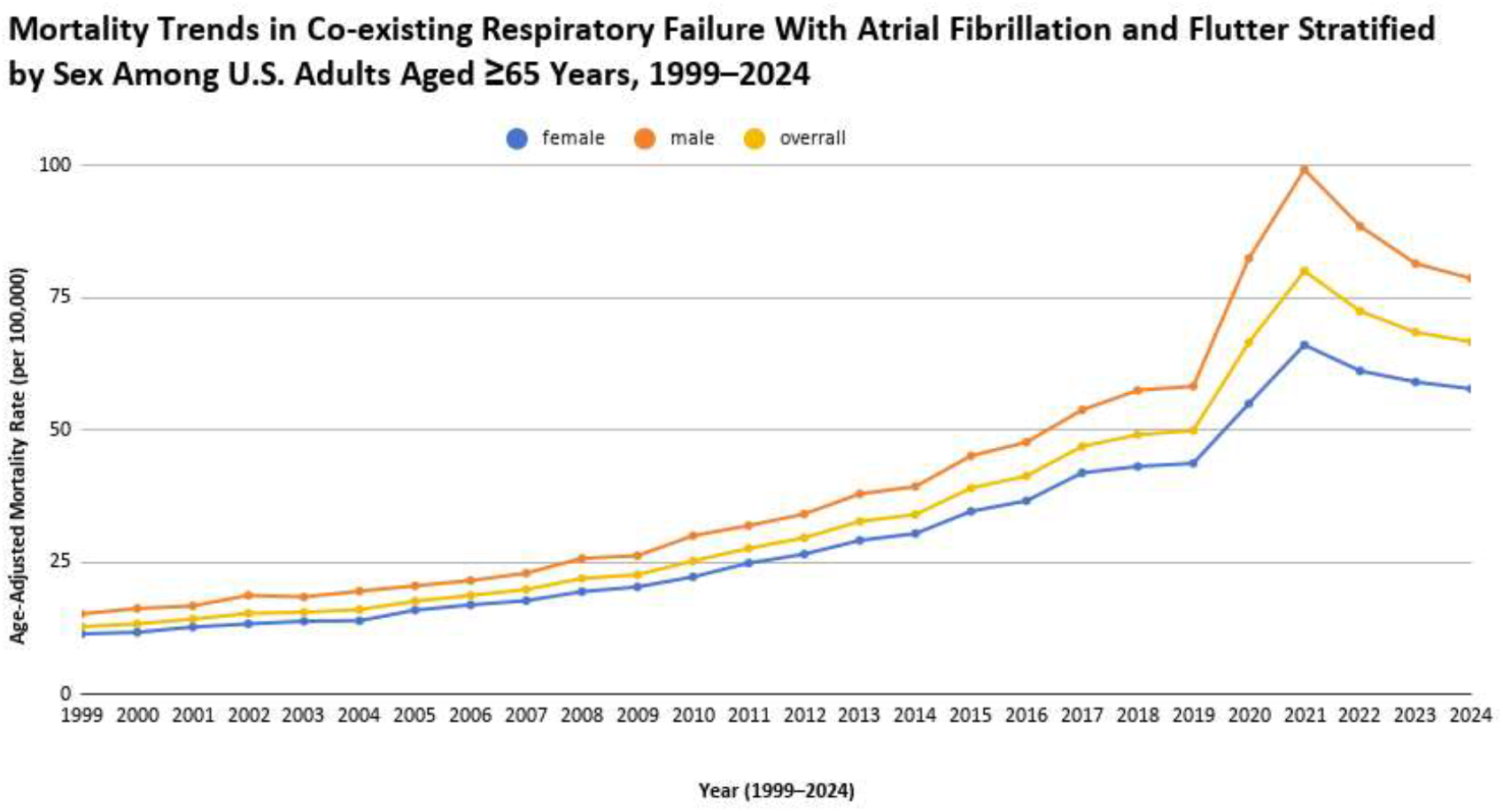
Mortality Trends by Demographic Characteristics (B) Stratified by Overall Period and Sex.

**Table 1.** Average Annual Percentage Change (AAPC) in Mortality Trends Across Cohorts (1999–2024)

| Cohort | Period | AAPC (95% CI) | P-value | Test Statistic |
| --- | --- | --- | --- | --- |
| Gender |  |  |  |  |
| Female | 1999–2024 | 7.81 (7.23, 8.38) | < 0.001 | 29.10 |
| Male | 1999–2024 | 8.44 (7.47, 9.41) | < 0.001 | 18.72 |
| Overall | 1999–2024 | 8.11 (7.35, 8.87) | < 0.001 | 22.93 |
| Census Region |  |  |  |  |
| Northeast | 1999–2024 | 6.67 (6.03, 7.32) | < 0.001 | 22.06 |
| Midwest | 1999–2024 | 9.35 (8.42, 10.30) | < 0.001 | 21.48 |
| South | 1999–2024 | 8.49 (7.70, 9.29) | < 0.001 | 22.94 |
| West | 1999–2024 | 7.94 (7.16, 8.72) | < 0.001 | 21.85 |
| **Race/Ethnicity** |  |  |  |  |
| Hispanic or Latino | 1999–2024 | 8.72 (7.19, 10.26) | < 0.001 | 12.19 |
| NH Asian or Pacific Islander | 1999–2024 | 6.23 (5.13, 7.35) | < 0.001 | 11.95 |
| NH Black or African American | 1999–2024 | 8.58 (7.52, 9.64) | < 0.001 | 17.38 |
| NH White | 1999–2024 | 8.37 (7.69, 9.05) | < 0.001 | 26.29 |
| **Urbanization** |  |  |  |  |
| Metropolitan | 1999–2020* | 8.21 (7.63, 8.78) | < 0.001 | 30.92 |
| Non-Metropolitan | 1999–2020* | 10.09 (9.25, 10.94) | < 0.001 | 26.11 |

*Note: NH, Non-Hispanic; AAPC, Average Annual Percentage Change; CI, Confidence Interval.*
*\*Urbanization data ends in 2020 due to reporting endpoint constraints.*

### 3.3 Mortality Trends Stratified by Race and Ethnicity

NH White adults experienced the highest absolute mortality burden with a mean AAMR of 38.6. Conversely, Hispanic or Latino adults demonstrated the greatest relative overall annual increase in mortality (AAPC = 8.72%), followed closely by NH Black adults (AAPC = 8.58%), NH White adults (AAPC= 8.37%), while NH Asian/Pacific Islander adults showed the lowest rate of increase (AAPC = 6.23%).

Joinpoint segments revealed distinct racial/ethnic variations. For Hispanic or Latino adults, AAMRs rose during 1999–2018 (APC = 7.92%), accelerated during 2018–2021 (APC = 24.82%), and declined during 2021–2024 (APC = −12.40%). NH Black adults followed a similar pattern (1999–2018 APC = 7.29%; 2018–2021 APC = 23.57%; 2021–2024 APC = −6.68%). For NH White adults, AAMRs increased during 1999–2018 (APC = 7.93%), surged during 2018–2021 (APC = 17.86%), and fell during 2021–2024 (APC = −4.46%). NH Asian/Pacific Islander trends shifted slightly later, rising during 1999–2019 (APC = 5.05%), peaking during 2019–2022 (APC = 33.84%), and declining during 2022–2024 (APC = −20.50%).

**Figure 1b.**
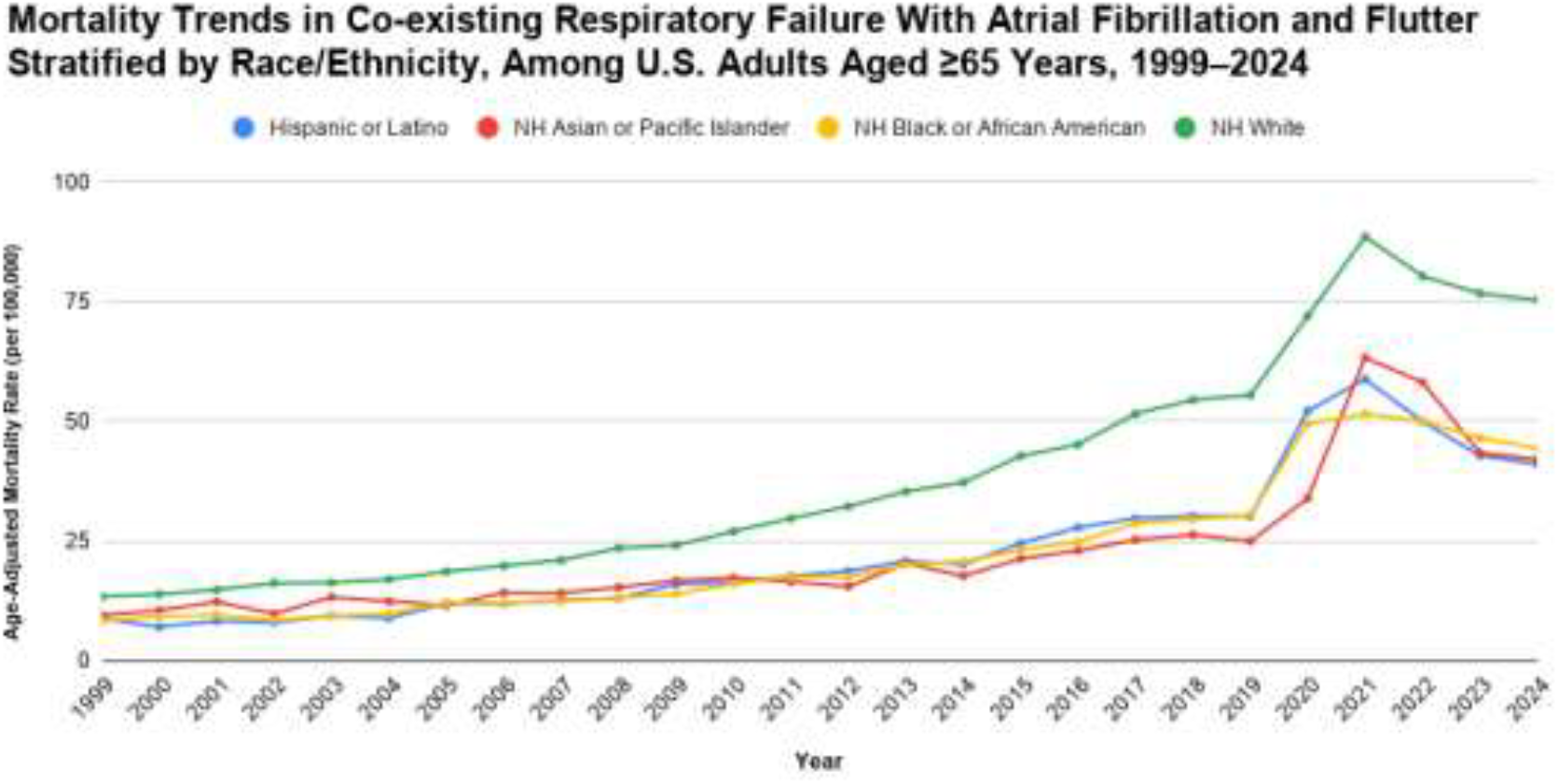
Mortality Trends by Demographic Characteristics (C) Race.

### 3.4 Mortality Trends Stratified by Place of Death

The vast majority of deaths occurred in institutional care settings, led by hospital inpatients (n = 277,815; 64.1%), followed by nursing homes/long-term care facilities (n = 61,120; 14.1%), the decedent’s home (n = 51,488; 11.9%), and hospice facilities (n = 22,818; 5.3%). Outpatient clinics, emergency departments, and other or unknown locations accounted for the remaining 4.6% of deaths.

**Figure 2.**
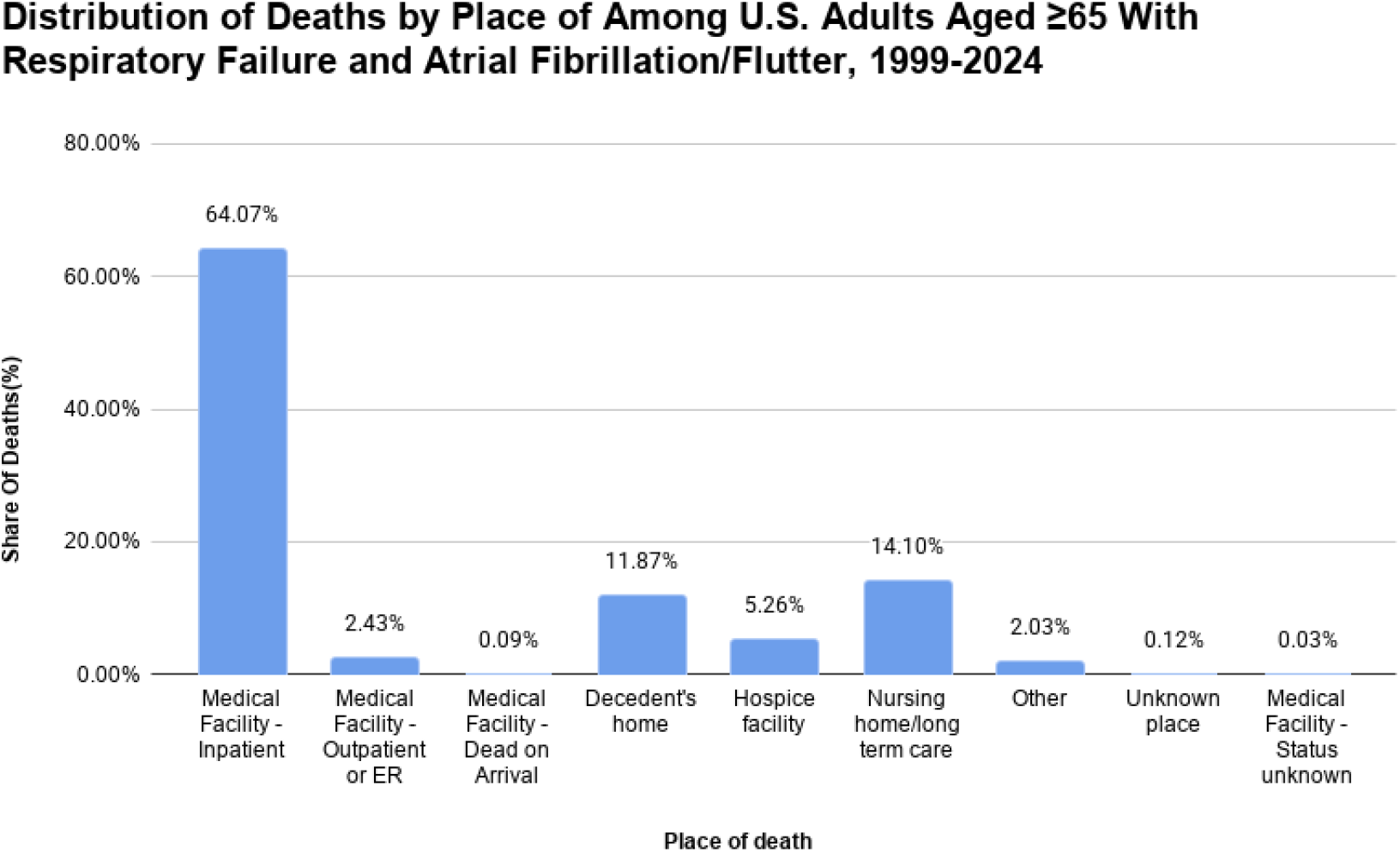
Mortality Patterns Stratified by Place of Death.

### 3.5 Mortality Trends Stratified by Census Region

All four US Census regions experienced rising mortality rates before 2021, followed by significant declines, with the highest AAPCs in Midwest (AAPC: 9.35%) and South (AAPC: 8.49%). In the Northeast, the AAMR increased from 14.6 in 1999 to 58.1 in 2024. AAMRs rose during 1999–2018 (APC = 6.28%) and 2018–2021 (APC = 15.32%) before declining during 2021–2024 (APC = −5.63%) and AAPC was 6.67%. The Midwest AAMR increased from 9.8 in 1999 to 65.2 in 2024. AAMR rose during 1999– 2018 (APC = 8.96%) and 2018–2021 (APC = 20.83%), followed by a sharp drop during 2021–2024 (APC = −6.82%). The AAMR in West increased from 16.3 to 77.9 with changes in significant APCs during 1999–2018 (APC = 7.13%) and 2018–2021 (APC = 18.47%) before falling during 2021–2024 (APC = −4.21%), and AAPC was 7.94%. In the South, AMMR South increased from 12 in 1999 to 65.1 in 2024 with three distinct periods of significant acceleration: 1999–2005 (APC = 4.53%), 2005–2018 (APC = 8.81%), and 2018–2021 (APC = 16.84%), followed by a post-2021 reversal (APC = −5.47%). *(Note: All regional trend values achieved statistical significance at P <* .*05)*.

**Figure 3.**
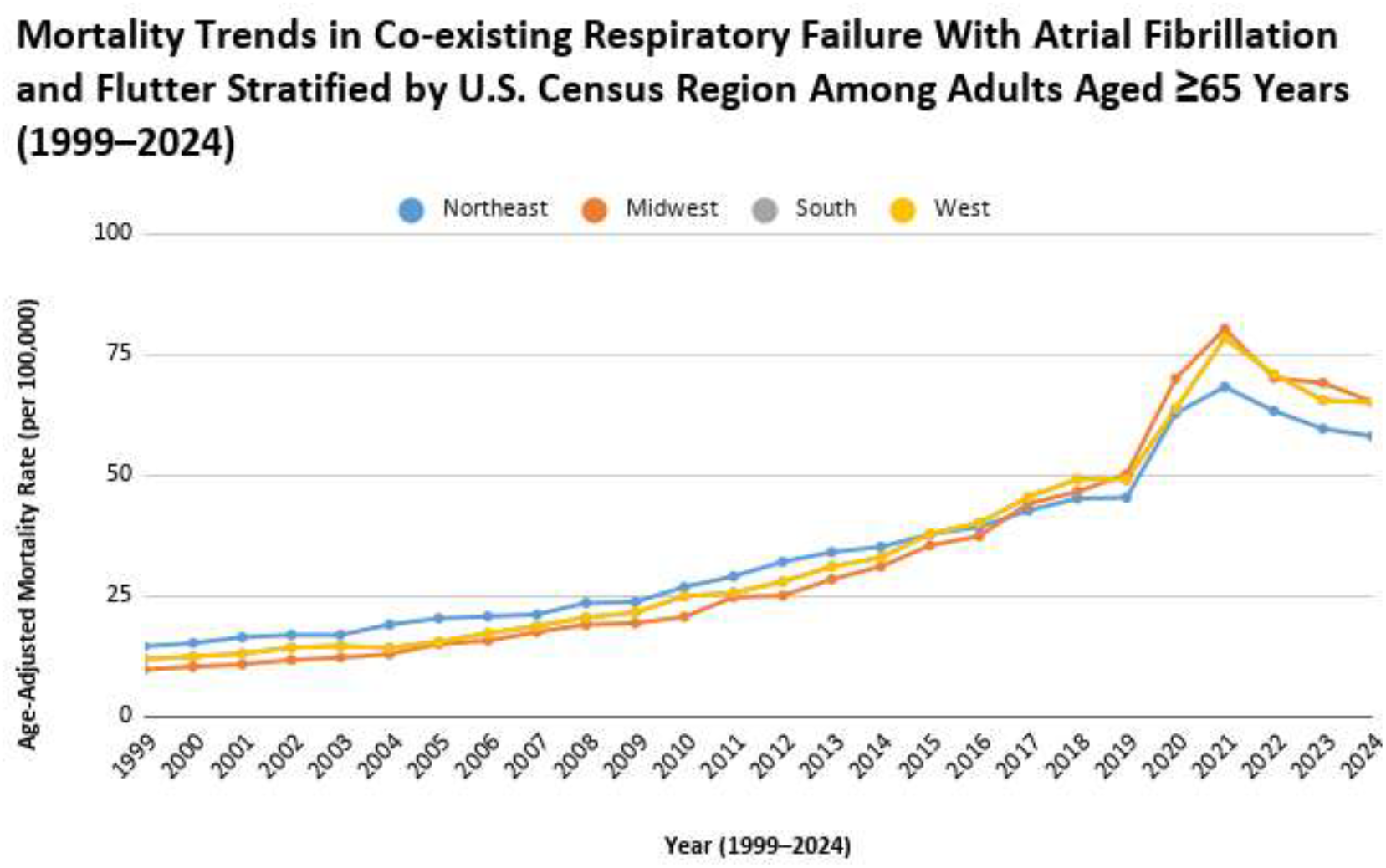
Mortality Patterns by Geographic Characteristics A. Census Region.

### 3.6 Mortality Trends Stratified by Urbanization

Due to variations in NCHS dataset transition formatting, urbanization-specific analysis was restricted to the 1999–2020 period. Mortality increased significantly across both settings, although non-metropolitan areas experienced a faster overall rate of increase (AAPC: 10.09% vs 8.21%; *P* < .0001). In metropolitan areas, the AAMR rose from 13.10 in 1999 to 66.43 per 100,000 in 2020, accelerating between 1999–2007 (APC = 5.26%) and 2007–2020 (APC = 9.11%). In non-metropolitan areas, the AAMR climbed from 11.05 in 1999 to 79.40 per 100,000 in 2020, driven by progressively faster compounding segments: 1999–2009 (APC = 6.71%), 2009–2018 (APC = 10.86%), and a stark 2018–2020 acceleration (APC = 20.57%).

**Figure 3a.**
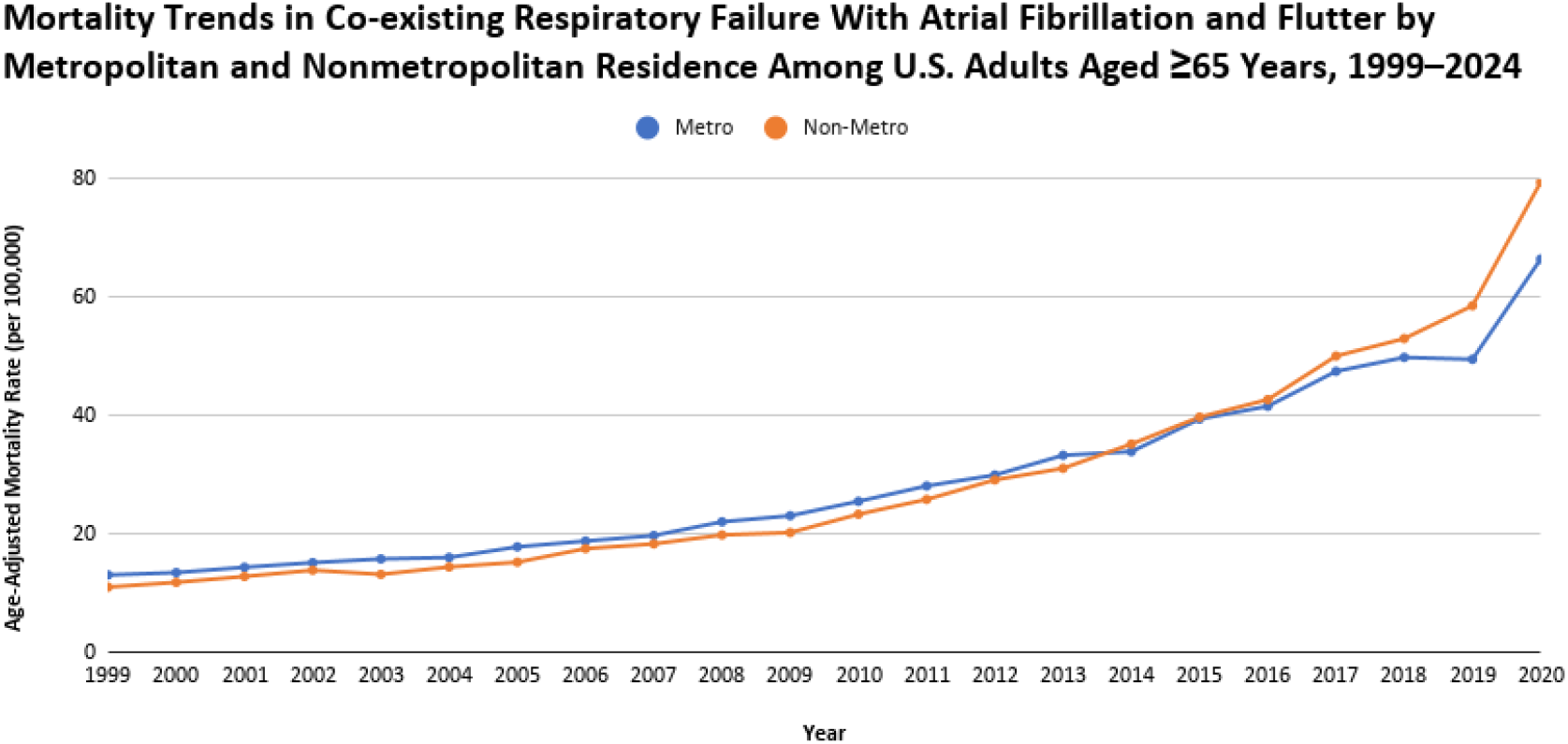
Mortality Patterns by Geographic Characteristics B. Urbanization.

### 3.7 Mortality Trends Stratified by State

State-level evaluations revealed substantial geographic variation. The lowest cumulative AAMRs (≤10th percentile) were clustered in Ohio (24.70), Hawaii (25.20), Alabama (27.80), West Virginia (29.60), Washington (31.00), and Pennsylvania (31.15) per 100,000 population. The highest mortality burdens (≥90th percentile) in terms of AAMRs were recorded in California (79.80), Rhode Island (79.80), Texas (80.95), Iowa (81.10), Virginia (82.90), and Louisiana (85.10).

**Figure 3b.**
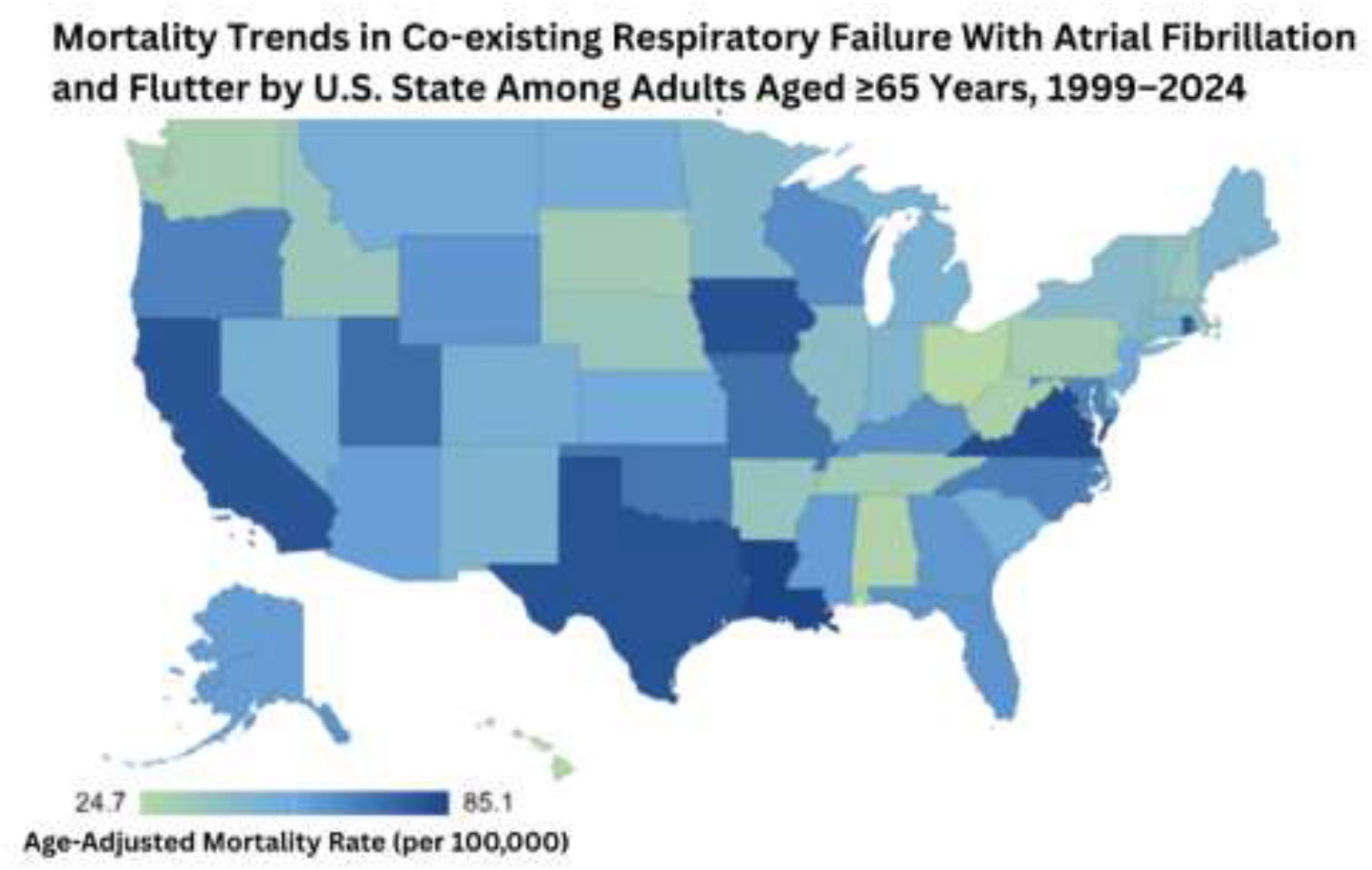
Mortality Patterns by Geographic Characteristics C. State.

## 4. Discussion

This nationwide CDC WONDER analysis demonstrates a substantial and sustained increase in mortality associated with concomitant respiratory (RF) and atrial fibrillation/flutter (AF) in older adults (65 years or above) in the United States from 1999 to 2024. The overall age adjusted mortality rate (AAMR) increased from 12.8 in 1999 to 66.7 in 2024. During this period there was gradual rise in AAMR (80.1 per 100,000) until 2021 followed by a partial decline (66.7 per 100,000) until 2024. The Mortality burden impacted males more in older adults and AAMR was highest (37.54) in Non-Hispanic (NH) White people while it was lower in Non-Hispanic Asian or Pacific Islander (22.03). State levels AAMRs ranged from 24.7 in Ohio to 85.1 in Louisiana. Lastly, COVID-19 pandemic impacted had a significant impact, with a shocking rise in AAMR to 80.1 per 100,000 (by 2021, peaking during the COVID-19 pandemic.

A study by Shu et al. reported AAMR for respiratory failure among adults aged ≥45 years increased from 3.71 in 1999 to 10.50 in 2023. This study also elaborated that overall AAPC was 4.67% in the United States^(1)^. Our findings extend this evidence by examining respiratory failure mortality specifically in the presence of atrial fibrillation/flutter among adults aged ≥65 years. Previous clinical studies have reported higher mortality among patients with respiratory failure who also have AF. In our study, analysis shows that mortality involving co-listed respiratory failure and AF/AFL among U.S. adults aged ≥65 years increased manyfold from 1999–2024.

A study conducted in 2024 regarding intensive care patients, with acute respiratory failure, showed that the presence of AF was associated with increased mortality during the COVID-19 pandemic period^(15)^. Our analysis also highlighted spike in AAMR during this time period. A 2025 study of patients with hypercapnic type 2 respiratory failure depicted that AF was associated with shorter survival and a 1.6-fold higher mortality risk as compared to patients without AF^(16)^. A recent MIMIC-IV/MIMIC-III study developed and externally validated prediction models for AF among patients with acute respiratory failure. In this study age was identified as the most important predictor of AF risk. These findings support the clinical relevance of examining AF/AFL in older adults with respiratory failure^(17)^.

The coexistence of respiratory failure and AF/AFL may reflect interlinked mechanisms which include hypoxemia, hypercapnia, autonomic imbalance, and inflammation. Moreover oxidative stress and atrial remodeling contribute to the worsening of condition. Respiratory dysfunction may also promote pulmonary hypertension which increases right-sided cardiac pressure and atrial stretch. These mechanisms may increase susceptibility to atrial arrhythmias^(18,19,)^. A Rapid ventricular response occurs during AF/AFL which can precipitate acute cardiogenic pulmonary edema, a contributing factor to acute respiratory failure through mechanism of pulmonary congestion and impaired gas exchange^(7)^.

AAMR involving respiratory failure and AF/AFL increased progressively from 1999 to 2018, potentially reflecting increased respiratory and cardiovascular disease burden in an aging population. The same pattern was observed in U.S. studies indicating increasing respiratory failure incidence during 2002–2017 and a higher number of deaths due to acute respiratory failure during 2014–2018^(20,21)^. The sharp rise in AAMR during COVID-19 pandemic from 2018 to 2021, peaking in 2021, which was associated with respiratory failure, atrial fibrillation, increased mortality, and greater mechanical ventilation requirements. Studies also showed increased the burden of atrial arrhythmias, particularly among older adults^(22,23)^. This rise also reflects substantial healthcare-system stress and impediment in access to critical care, particularly for patients requiring complex respiratory and cardiovascular management^(24)^.

In our study, AAMR declined during the post-pandemic period, which was consistent with U.S. data showing reduction in age-adjusted COVID-19 mortality from 2021 to 2022 and further decline in 2023^(25,26)^. A separate CDC WONDER analysis similarly reported declining AF-related mortality after the pandemic peak^(27)^.

The post-pandemic decline in mortality may be attributed to advent COVID-19 vaccination which helped increase immunity in the population^(28)^, the emergence of less fatal variants (such as Omicron)^(29)^, and improved management utilizing antiviral and immunomodulatory therapies. Despite the post-pandemic decline, mortality rates remained substantially higher than early-2000s levels. Similar declines after the pandemic peak have been reported for COVID-19 mortality, AF-related mortality, and mortality involving respiratory failure with chronic kidney disease^(30)^, suggesting that the pandemic may have amplified preexisting vulnerabilities rather than representing only a transient mortality shock.

There is significant demographic and geographic heterogeneity in mortality involving co-existing respiratory failure and atrial fibrillation/flutter among U.S. adults aged ≥65 years. Despite the fact males and females showed similar overall temporal trajectories, males had a greater long-term increase in mortality and a greater rise during 2018–2021, followed by a more pronounced decline after 2021. This pattern matches with previous mortality analyses reporting higher atrial fibrillation-related and respiratory failure-related mortality among men, showing that prevalence of these diseases is higher in men^(1,31)^. Differences in the burden of cardiopulmonary ailments, underlying risk factors, healthcare services and disease severity may contribute to the observed sex differences.

Racial and ethnic disparities were evident in mortality involving co-existing respiratory failure and atrial fibrillation/flutter. Non Hispanic (NH) White adults experienced the highest mortality burden. Transportation issues, lack of specialty services, rural hospital closures and delays in emergency responses are the reasons for worse consequences of respiratory failure and atrial fibrillation/flutter^(32,33)^.

Similar racial variations among NH White has been reported in U.S. mortality studies involving atrial fibrillation and cardiopulmonary conditions. These differences indicate variations in cardiopulmonary comorbidity burden^(34)^. Asian or Pacific Islander adults showed the lowest overall rate of increase; this may reflect differences in smoking prevalence, less deranged cardiovascular profiles, stronger social support and decreased prevalence of chronic respiratory diseases^(35,36)^.

However, these factors could not be directly evaluated in CDC WONDER. There exists pandemic era heterogeneity as race has largest pandemic-era APC rise which can be attributed to lack of instant testing access and delays in presenting to hospitals in accessibility, and socioeconomic status also contributed to these results^(38)^.

The West had the highest AVERAGE AAMR (40.9 per 100,000), consistent with prior evidence of higher AF-associated cardiopulmonary mortality in the West^(39)^. Regional differences in risk factors and healthcare access may contribute. In our study, an urban–rural disparity was observed, with mortality consistently higher in non-metropolitan than metropolitan areas during 1999–2020. A marked elevation was seen during 2018–2020. Subgroup analysis further demonstrated that this acceleration was most prominent in Non-Core rural areas. Previous studies have described increasing rural hospital closures in the United States, which may reduce local healthcare capacity^(32)^. Rural hospital closures have also been associated with longer emergency medical transport and total activation times^(33)^Collectively, these findings suggest that differences in healthcare access and emergency care infrastructure may contribute to the higher mortality observed in non-metropolitan areas. The higher number of deaths in medical facilities followed by nursing homes indicates huge strain of this comorbidity on healthcare setups^(40)^.

This analysis infers the rise in mortality highlights the susceptibility of this population during periods of severe respiratory and cardiovascular stress. Consequently, reducing this mortality burden requires a highly integrated approach that recognizes respiratory failure and AF/AFL as interconnected clinical conditions rather than isolated organ-specific disorders. This approach must emphasize the early identification of arrhythmias. By optimizing respiratory function, managing cardiovascular risk factors, and coordinating cardiopulmonary care, better results can be obtained for patients.

## 5. Limitations

This study has several limitations which inherent to large-database observational analyses. First, data tracking relies strictly on ICD-10 administrative coding from death certificates via the CDC WONDER system. It introduces potential risks of misclassification, incomplete documentation, or underreporting of secondary comorbidities like AF/AFL. Second, the dataset lacks granular clinical, laboratory, and pharmacological variables. Consequently, we could not evaluate crucial survival determinants such as specific respiratory failure phenotypes (Type 1 vs. Type 2), echocardiographic metrics, or the use of mechanical ventilation strategies, and cardiopulmonary medications (e.g., anticoagulants and antiarrhythmics). Because of the ways atrial fibrillation and flutter are coded in database, they could not be analyzed separately. Third, certain demographic populations, specifically American Indian or Alaska Native individuals, were excluded because of data unreliability thresholds within CDC WONDER. Individuals with multiple races could not be tracked consistently across the entire 1999–2024 period due to temporal shifts in database subcategories. Lastly, given the retrospective design of this trend analysis, these findings demonstrate substantial population-level associations but cannot be used to deduce direct, patient-level clinical causality.

## Data Availability

All data produced are available online at (https://www.google.com/search?q=CDC+wonder+analysis&ie=UTF-8)

